# Is the Current N95 Respirator Filtration Efficiency Test Sufficient for Evaluating Protection Against Submicrometer Particles Containing SARS-CoV-2?

**DOI:** 10.1101/2020.05.14.20102327

**Authors:** Changjie Cai, Evan L. Floyd, Kathleen A. Aithinne, Toluwanimi Oni

## Abstract

The National Institute of Occupational Safety and Health procedure No. TEB-APR-STP-0059 recommend of measuring the respirator filtration efficiency using sodium chloride aerosol with count median diameter of 75 nm ± 20 nm and geometric standard deviation ≤1.86. This study showed that this method would overestimate the respirators’ ability to protect against submicrometer particles. In this study, we converted both mobility diameter and equivalent volume diameter to aerodynamic diameter for comparison. The results showed that one unqualified KN95 respirator (with the filtration efficiency of 72%±3% for ≥300 nm sodium chloride aerosol) still passed the test with a measured overall filtration efficiency of 98%±3%, due to its larger most penetrating particle size compared to the typical N95 respirator. In addition, after three cycle H_2_O_2_ plasma vaporous sterilizations, the most penetrating particle size for the N95 grade respirators also shifted to 250 nm – 500 nm, in which size the particles carried the peak concentration of the SARS-CoV-2 in hospitals. This size shift caused the significant difference between the size specific (250 nm – 500 nm) filtration efficiency and overall filtration efficiency using the same NaCl test aerosol. For example, after three cycle H_2_O_2_ plasma vaporous sterilizations, the size specific filtration efficiency of the N95 was 55%±2%, however, the measured overall filtration efficiency was still 86%±5%. The size Specific filtration efficiency of the KN95 was 69%±2%, but, the measured overall filtration efficiency was still 90%±3%. In order to protect health care personnel adequately, we recommend increasing the test aerosol size, and measuring the size specific filtration efficiency to evaluate the N95 alternatives (e.g. KN95), and the reuse of N95 level respirators. In addition, multi-cycle sterilization with ultraviolet germicidal irradiation appears to have fewer negative effects than H_2_O_2_.

## 1. Introduction

Recent studies indicates that aerosol transmission of the severe acute respiratory syndrome coronavirus 2 (SARS-CoV-2) is plausible since the virus can remain viable and infectious in aerosol form for hours (*1,2*). Wearing personal protective equipment (e.g. N95 respirator) could be an effective strategy before the effective vaccines or antiviral drugs. The N95 grade respirator should have a minimum filtration efficiency of 95% for ≥300 nm (aerodynamic diameter) sodium chloride (NaCl) aerosols. Due to the COVID-19 pandemic and shortage of mask supplies (*3*), some studies analyzed the reuse of N95/KN95 respirators after sterilization by measuring the filtration efficiency using NaCl test aerosol (*4,5*). However, in the U.S. Centers for Disease Control and Prevention (CDC) National Institute of Occupational Safety and Health (NIOSH) procedure No. TEB-APR-STP-0059, the count median diameter (CMD) of the NaCl test aerosol is 75 nm ± 20 nm with the geometric standard deviation (GSD) ≤1.86, which might be too small compared to the particles containing infectious agents.

The peak concentration of the SARS-CoV-2 in hospitals was found in 250 nm – 500 nm (aerodynamic diameter) particles (*6*), although the actual SARS-CoV-2 size ranges only from 60 nm – 150 nm (*7,8,9*). Since the most penetrating particle size (MPPS) for typical N95 respirators is 30 nm – 100 nm (*10*), the CDC NIOSH recommends using the NaCl test aerosol with a CMD of 75 nm ± 20 nm and GSD of ≤1.86. However, using this size NaCl test aerosol means that around 89.9% particles are ≤250 nm (aerodynamic diameter), which is not in the particle size range of containing the peak concentration of the SARS-CoV-2 in hospitals. The light-scattering instruments (e.g., aerosol photometer), which are commonly used for filtration efficiency test, are usually limited to measuring aerosol larger than 100 nm (*11*); still around 84.6% particles are ≤250 nm (aerodynamic diameter) if a photometer is used during the test. Therefore, a respirator efficient for particles ≤250 nm may not capture most SARS-CoV-2, and the size specific filtration efficiency is a better assessment of the respirator’s protection ability rather than the overall filtration efficiency to prevent the SARS-CoV-2 transmit via airborne particles.

The goal of this demonstration study was to compare the size specific filtration efficiency to the overall filtration efficiency through testing an untreated (new) KN95, and treated (multiple sterilization cycles) KN95/N95 respirators.

## 2. Methods

### 2.1 Experimental Setup

We tested KN95 respirators from over 20 various manufacturers, and selected two of them (one qualified and one unqualified) for analysis in this study. We analyzed the reasons why the unqualified KN95 respirator (Nine Particles Medical Equipment Co, Ltd, Yongzhou, Hunan, China) still passed the test using the NIOSH recommended NaCl test aerosol. We also compared the effects of two- and three-cycle sterilization using vaporous plasma hydrogen peroxide (H_2_O_2_) and ultraviolet (UV) germicidal irradiation on the filtration efficiencies by size of KN95 (qualified one from Civilian Antivirus, Qingdao, Shandong, China) and N95 (model 1860, 3M, St Paul, MN, USA) respirator. The experimental setup was illustrated in previous study (*12*). A NaCl aerosol was generated using a Collision Nebulizer (3 jet, CH Technologies, Westwood, NJ, USA) operated at 20 psi using 2% NaCl aerosol recommended by NIOSH procedure No. TEB-APR-STP-0059. We tested five samples for each respirator. A scanning mobility particle sizer (SMPS, model 3936, TSI Inc., Shoreview, MN, USA) was used to measure the number concentration of test aerosol from 16.8 nm to 514 nm up and down stream of each sample. We conducted a two-sample t-test (with *α=0.05* as significance threshold) to compare the mean filtration efficiency of the five samples at each aerosol size between two and three sterilization cycles.

### 2.2 Diameter Conversion

In this study, we converted both mobility diameter and equivalent volume diameter to aerodynamic diameter for comparison. The equations are from literature (*13, 14*):

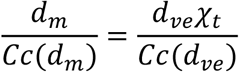

where *d_m_* and *d_ve_* are mobility diameter and volume equivalent diameter, respectively; *Cc(d_m_)* and *Cc(d_ve_)* are the respective slip correction factors for *d_m_* and *d_ve_*; *χ_t_* is the dynamic shape factor in transition regime.

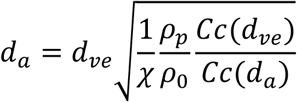

where *d_a_* is aerodynamic diameter; *ρ_p_* and *ρ_0_* are particle density and standard density (1g cm^-3^), respective; *Cc(d_a_)* is the slip correction factors for *d_a_*; *χ* is the dynamic shape factor. The salt aerosol density is 1.75 – 1.99 g cm^-3^, and the dynamic shape factor is 1.05 – 1.14 from measurements (*15*). Therefore, we assumed the salt aerosol density of 1.8 g cm^-3^ and shape factor of 1.1 in this study.

## 3. Results

### 3.1 Effects of test aerosol size on filtration efficiency

We found an interesting case for an unqualified KN95 shown in **Figure 1**. The results showed that the respirator passed the filtration efficiency test with an overall filtration efficiency of 98±3%, and an acceptable pressure drop of 0.173 “wg ± 0.015 “wg (**Figure 1a**). However, by looking at the filtration efficiency by aerosol size, this respirator is not qualified as a N95 grade respirator since the filtration efficiency for particles ≥300 nm is 72%±3%, which is much lower than a minimum of 95% (**Figure 1b**).

**Figure 1.**
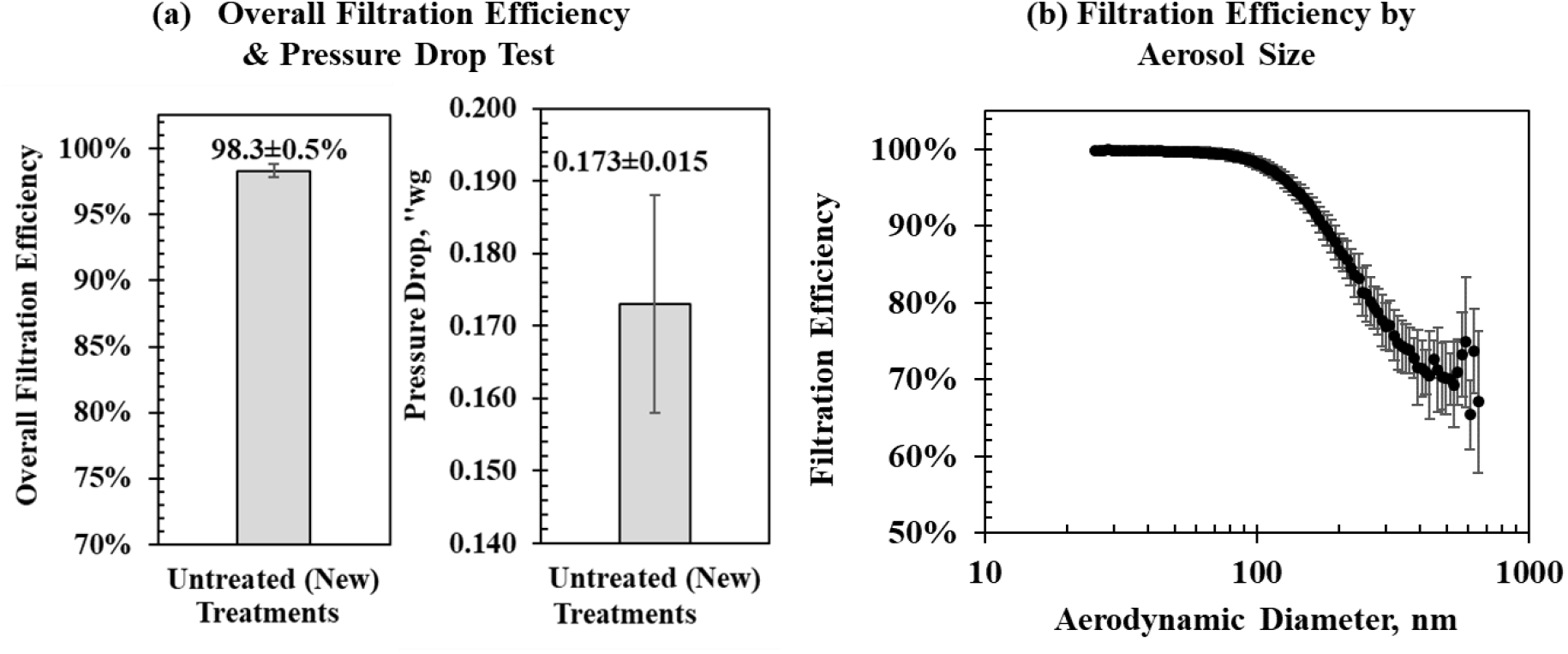
Test of an unqualified KN95 respirator: (a) overall filtration efficiency & pressure drop test, and (b) filtration efficiency by aerosol size.

We conducted a sensitivity study of estimating the overall filtration efficiency by increasing the CMD from 55 nm (79 nm equivalent count median aerodynamic diameter, CMAD) to 215 nm (278 nm equivalent CMAD) (**Figure 2**). The results indicated that the overall filtration efficiency would gradually decrease from 97% to 82% by increasing the test aerosol CMAD from 79 nm to 278 nm.

**Figure 2.**
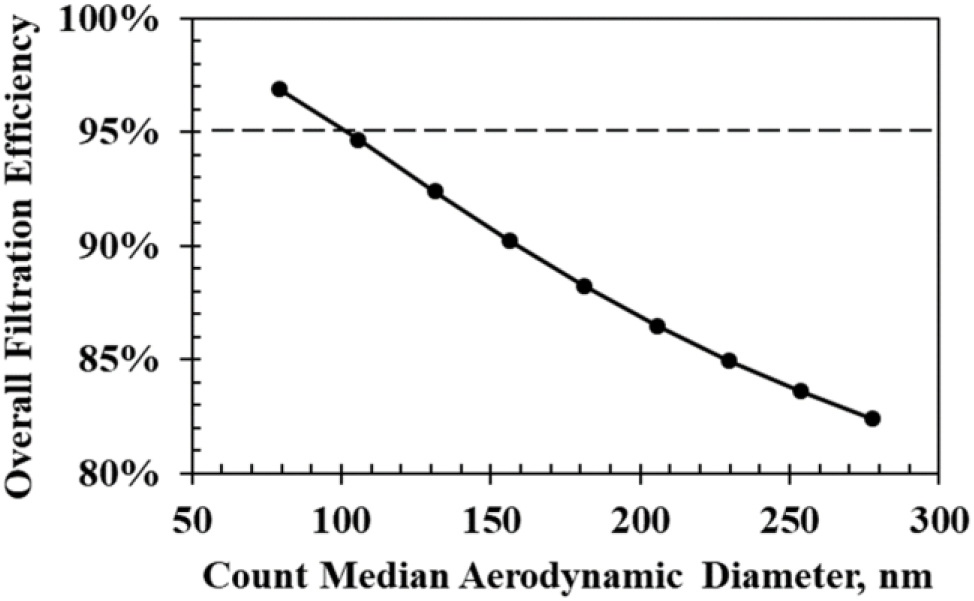
Effects of the test aerosol size on the overall filtration efficiency.

### 3.2 Effects of multi-cycle sterilization on the filtration efficiency

The effects of multi-cycle sterilization on the filtration efficiency by aerosol size are summarized in **Figure 3**. After three H_2_O_2_ sterilization cycles, the filtration efficiency for 250 nm – 500 nm particles dropped from 84%±2% to 69%±2% for KN95, and from 97%±2% to 55%±2% for N95. The differences between two and three cycles were significant for both KN95 (*p*-value = 0.014±0.014) and N95 (*p*-value = 0.0009±0.0008). After UV sterilization, the filtration efficiency of KN95 for 250 nm – 500 nm particles was 94.4%±1.0% after two cycles, and 95.0%±0.6% after three cycles; and the filtration efficiency of N95 was 95.9%±1.4% after two cycles, and 96.9%±0.7% after three cycles. The differences were not statistically significant between two and three cycles for both KN95 (*p*-value = 0.852±0.093) and N95 (*p*-value = 0.448±0.267).

**Figure 3.**
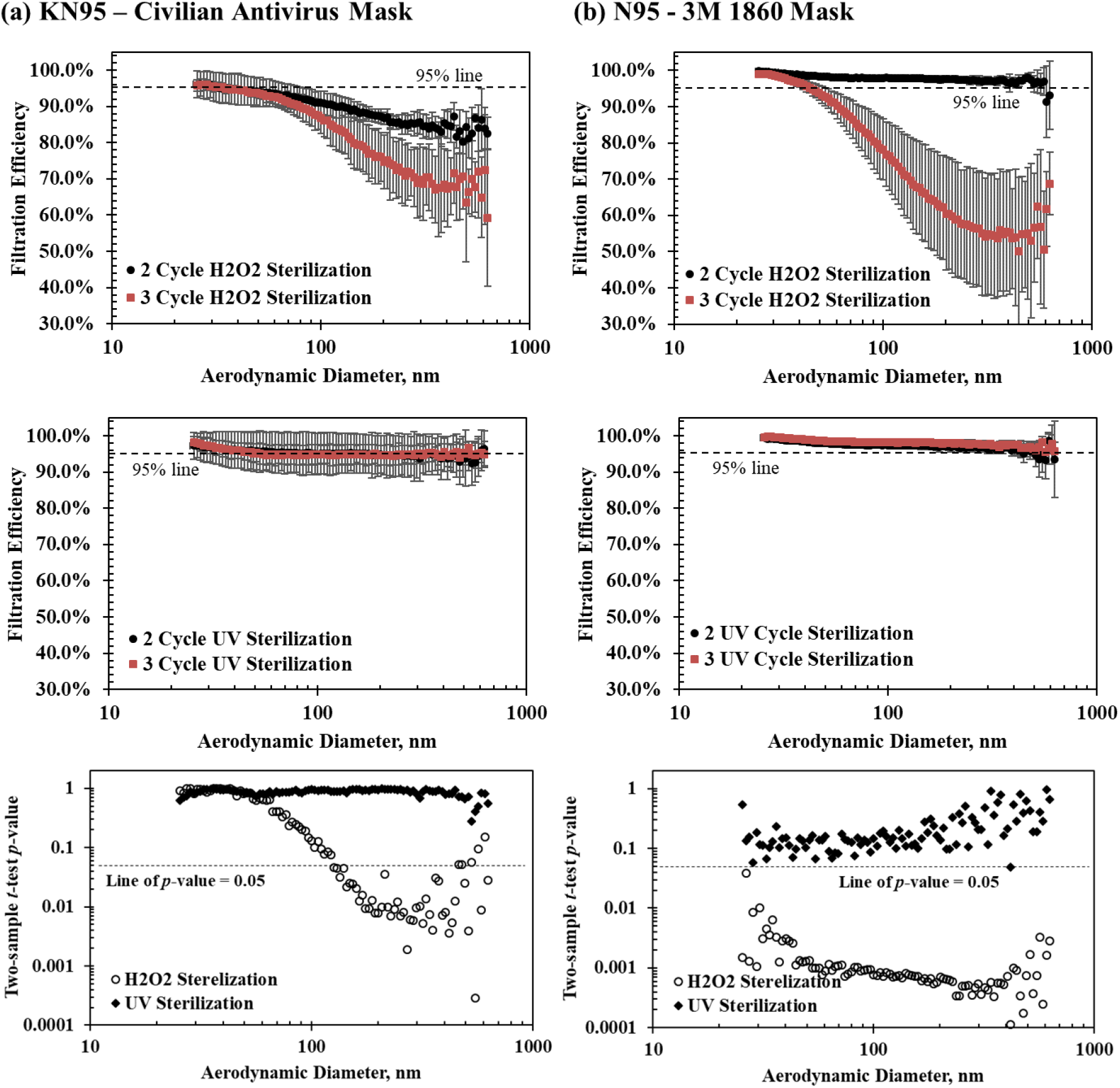
Effects of multi-cycle sterilization on filtration efficiency by aerosol size using H_2_O_2_ and UV treatment for (a) N95, and (b) KN9.

The size specific filtration efficiency is quite different from the overall filtration efficiency, especially after three cycle H_2_O_2_ treatment (see **Table 1**). For instance, the overall filtration efficiencies for KN95 and N95 after three cycle H_2_O_2_ treatment are 90%±3% and 86%±5%, respectively; however, the size specific filtration efficiencies are only 69%±2% for KN95 and 55%±2% for N95, respectively. The big difference between the size specific filtration efficiency and the overall filtration efficiency might be due to the MPPS shift after three cycle H_2_O_2_ treatment. The MPPS shifted from nanoparticle sizes (see **Figure 4**) to submicrometer particle sizes (see **Figure 3**), which are not the majority of the NaCl test aerosol size.

**Table 1.** Comparison of overall filtration efficiency and size specific (250 nm – 500 nm) filtration efficiency after multi-cycle H_2_O_2_ treatment.

| Respirator Type | H<sub>2</sub>O<sub>2</sub> Treatment | Overall Filtration Efficiency | Size Specific Filtration Efficiency |
| --- | --- | --- | --- |
| KN95 – Civilian Antivirus | 2 Cycle | 93%±3% | 84%±1% |
|  | 3 Cycle | 90%±3% | 69%±2% |
| N95 – 3M 1860 | 2 Cycle | 98%±1% | 97%±1% |
|  | 3 Cycle | 86%±5% | 55%±2% |

**Figure 4.**
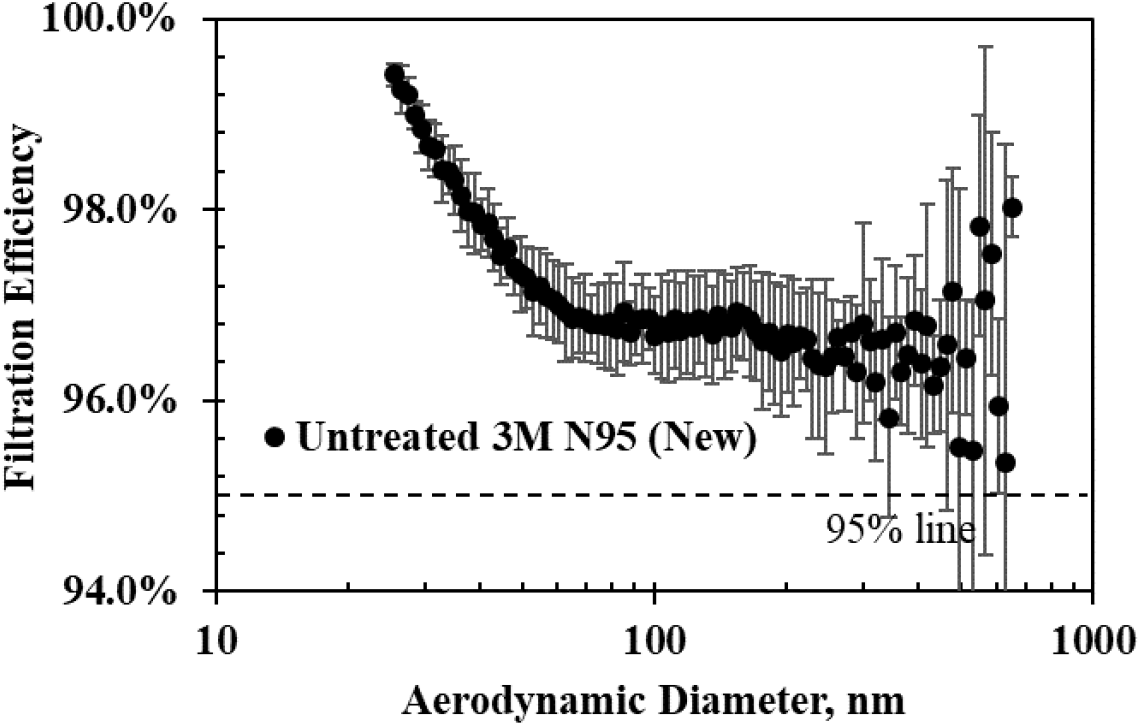
The most penetrating particle size (MPPS) for an untreated 3M N95 (new)

## 4. Discussion

This study proposed that the current NaCl test aerosol size might need to be increased for testing the new KN95 respirator, and the reuse of N95 level respirators after sterilizations due to their much larger MPPS compared to traditional N95 respirators’. In addition to considering the overall filtration efficiency, the filtration efficiencies for particle sizes similar to infectious particles should be considered. This study also found that the two different multi-cycle sterilization processes have unique effects on the filtration efficiencies by aerosol size of different respirators. Multi-cycle sterilization with UV appears to have fewer negative effects than H_2_O_2_. Limitations include the small variety of respirator manufacturers and the limited numbers of samples (n=5) for each respirator and only two sterilization techniques evaluated. In addition, this study only evaluated the filtration efficiency after three sterilization cycles as this corresponds with guidance from the American College of Surgeons for H_2_O_2_ sterilization. The filter material might degrade further with more cycles, which should be investigated for UV treatment.

## Data Availability

All data referred to in the manuscript is available.

## Acknowledgement

This work was made possible by support from the Oklahoma State Department of Health to establish this testing program, by the supply chain logistics group at the University of Oklahoma Medical Center who provided the respirators and sterilization treatments, by the University of Oklahoma Health Sciences Center VPR’s office through a COVID19 Rapid Response pilot grant, and by the student volunteers that assisted with data collection.

